# The significance of molecular heterogeneity in breast cancer batch correction and dataset integration

**DOI:** 10.1101/2024.12.22.24319524

**Authors:** Nicholas Moir, Dominic A. Pearce, Simon P. Langdon, T. Ian Simpson

## Abstract

Breast cancer research benefits from a substantial collection of gene expression datasets that are commonly integrated to increase analytical power. Gene expression batch effects arising between experimental batches, where signal differences confound true biological variation, must be addressed when integrating datasets and several approaches exist to address these technical differences. This brief communication study clearly demonstrates that popular batch correction techniques can significantly distort key biomarker expression signals. Through the implementation of ComBat batch correction and evaluation of integrated expression values, we profile the extent of these distortions and consider an additional mitigatory batch correction step. We demonstrate that leveraging *a priori* knowledge of sample molecular subtype classification can optimally remove batch effect distortion while preserving key biomarker expression variation and transcriptional legitimacy. To the best of our knowledge, this study presents the first analysis of the interplay between dataset molecular composition and the concomitant robustness of integrated, batch-corrected biological expression signal.

## Main text

The reliability and robustness of oncology gene expression profiling studies are affected by the size and quality of available sample data. Sample count has a major impact on the reliability of clinical gene expression analyses, yet the size of most previous studies has been driven by sample availability and cost. Nonetheless, enhanced biological insight can be empowered by integrating discrete datasets into a larger meta-dataset. The improved statistical power of downstream integrated analysis has been demonstrated^1^–6 and leveraged to identify and categorise molecular events within both breast cancer^7^ and the wider oncological research landscape^7^–10 that may be miscategorised or undetectable in smaller datasets. Direct integration of probe or transcript-level expression data from multiple studies is therefore potentially very powerful but technical batch-effects, manifesting both within and between studies, must be addressed before initiating integrated analysis^1^–3,11–14. Several groups have investigated optimal batch correction approaches, where expression data are augmented to remove batch effects while ensuring the maximal retention of signals representative of true biological variation. To this end, ourselves and others have previously stated that breast cancer datasets should only be integrated where they are suitably ‘similar’^2^, but enhanced understanding of these fundamental issues still appear elusive many years later. Additionally, we have cautioned that the molecular composition of breast cancer expression data does not accurately reflect breast cancer heterogeneity at the population level^15^, and have previously highlighted that integrating datasets with vastly varying molecular compositions can dramatically reduce the accuracy of prognosis predictions^2^.

Batch-effect correction has gained acceptance as a necessary dataset integration step but there remains little focus on the potential interplay between sample molecular subtype composition and batch correction robustness. In this study, we explore the effect of ComBat^16^ batch correction on fidelity of key breast cancer biomarker expression signals, and show that consideration of molecular heterogeneity is required to ensure the optimal preservation of vital gene expression signal integrity.

METABRIC data were collected between 1977 and 2005 from five centres in the UK and Canada and packaged as two stand-alone discovery (*n=997*) and validation *(n=995)*^17^ annotated expression datasets. The potential integration of these into a single 1992 sample dataset is of obvious analytical benefit in downstream molecular subtype and novel biomarker discovery pipeline workflows^18,19^. However, the existence of strong batch effects between the discovery and validation datasets has been cautioned against in published studies^18,20,21^.

As described, effective batch correction removes technical differences without undesirable further modification of the biological signal. In this brief communication, we evaluate the performance of several different ComBat implementations on expression values of the ILMN_1678535 (ESR1)^22^, ILMN_2352131 (ERBB2)^23^and ILMN_1680955 (AURKA)^24^ probes. These genes comprise the SCMGENE subtype classification model^25^ and are recognised as key breast cancer biomarkers of hormonal signalling and proliferation^25,26^. To investigate and illustrate the augmentation of these expression values during batch correction, we compare biomarker expression within the canonical PAM50 molecular subtypes (Basal, Luminal A, Luminal B, Her2+ and Normal Breast-like) between METABRIC discovery and validation datasets. Significant expression differences are expected between uncorrected data given the acknowledged presence of batch effects and, recognising sample variation as a sum of technical and biological differences, insight into the relative effectiveness of each ComBat approach is gained by comparing post-correction expression within each molecular subtype class. Substantial inter-sample molecular differences are encapsulated within PAM50 subtype assignment, and minimisation of significant variation within subtype classes therefore acts as an appropriate proxy for assessing and comparing batch correction efficacy.

In this study, we compare the effect of four different ComBat implementations on biomarker expression. ComBat implements a location-scale (LS) method, which models expression mean and variance within each batch, before adjusting expression according to these models. Default parametric correction assumes that the location batch effect variables originate from the same normal distribution and scale effect variables from the same inverse gamma distribution. Non-parametric correction, which uses a Monte Carlo integration-based technique to estimate the location and scale batch effect parameters^27^ is also evaluated. In addition to these parametric and non-parametric corrections, we include two approaches that account for dataset molecular heterogeneity by leveraging *a priori* PAM50 subtype assignment. We investigate including PAM50 molecular subtype as a defined covariate in the model used by ComBat and additionally introduce a further approach that leverages a pre-correction subtype stratification of samples. Subtype-stratified batch correction involves pooling samples of each molecular subtype from each batch and correcting each PAM50 molecular subtype group separately.

Transcriptome biomarker fidelity, represented as a minimisation of within-subtype sample expression difference following correction, is clearly enhanced when sample molecular heterogeneity is considered during batch correction. Statistically significant (two-sided Mann-Whitney-Wilcoxon test followed by Holm-Bonferroni correction) Aurora Kinase A (AURKA) expression (figure 1A) differences persists in both Luminal A (parametric p=.001, non-parametric p=.001) and Luminal B (parametric p<.001, non-parametric p<.001) samples following both parametric and non-parametric ComBat correction. In contrast, both *a priori* PAM50 batch correction approaches remove technical differences and result in no significant population expression differences between discovery and validation sample groups. The benefit of leveraging molecular subtype assignment is further demonstrated with Oestrogen receptor alpha (ESR1) expression (figure 1B). Rather disturbingly, uncorrected basal samples display no significant population difference but have significant population expression differences introduced through parametric (p<.001), non-parametric (p<.001) and PAM50 covariate correction (p<.001). HER2 samples retain differences following all four parametric (p<.001), non-parametric (p<.001), PAM50 stratified (p=.019) and PAM50 covariate (p<.001) correction approaches. Within luminal A samples, parametric (p<.001) and non-parametric (p<.001) perform poorly, while both *a priori* PAM50 approaches remove significant ESR1 expression differences between sample populations. Similarly, both parametric (p<.001) and non-parametric (p<.001) fail to resolve population differences in Luminal B samples, with PAM50 stratified correction retaining statistical significance between batches (p=.019) and only PAM50 covariate correction removing statistical significance. Parametric (p<.001) and non-parametric (p<.001) correction additionally performs poorly with normal-like samples, with both PAM50-based approaches removing significant population differences, a prerequisite for accurate post-integration analysis.

**Figure 1.**
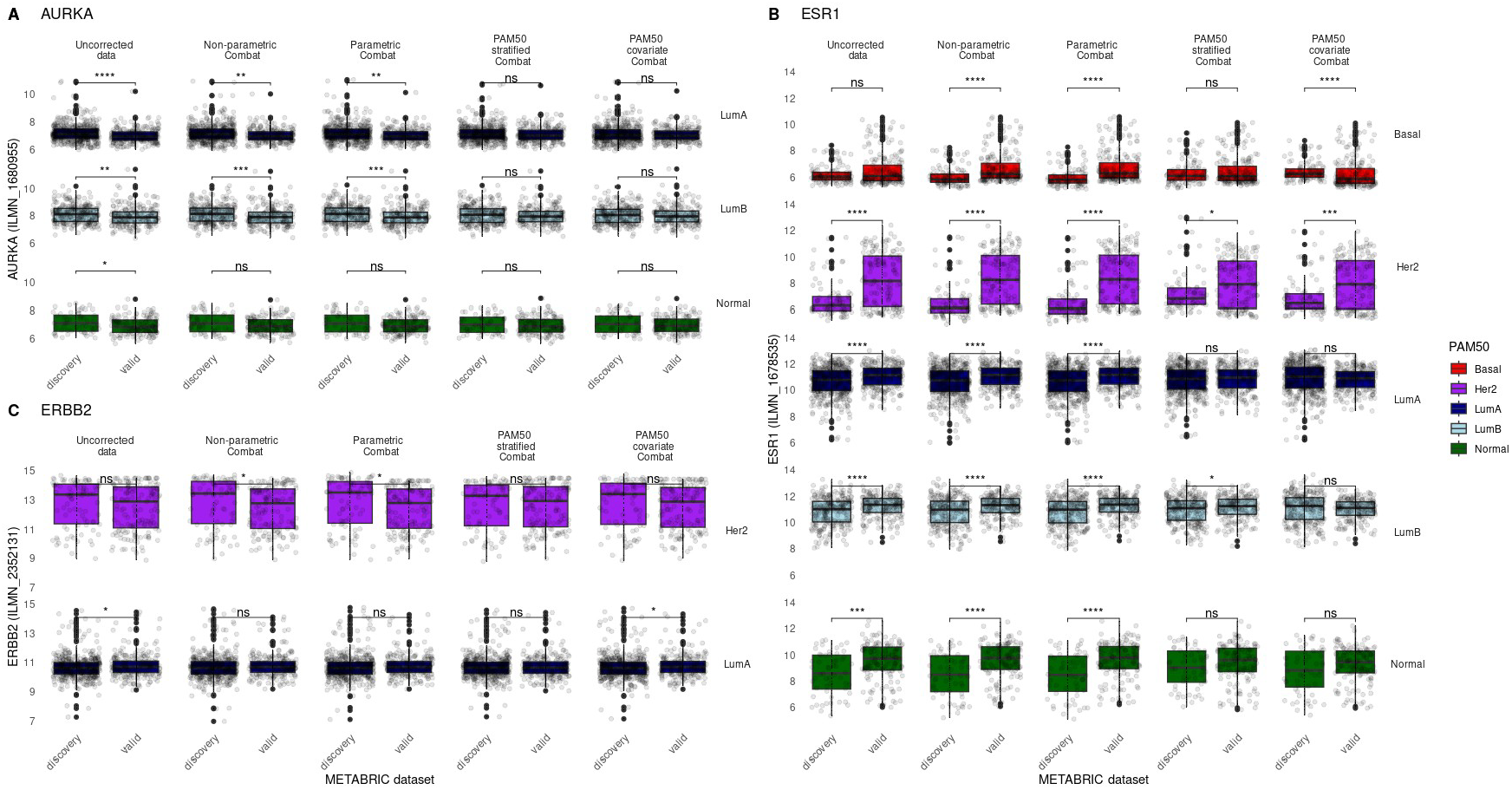
Effect of various ComBat batch correction implementations on METABRIC biomarker expression. Comparison of sample expression values before and after 5 batch corrections in AURKA (**A**), ESR1 (**B**) and ERBB2 (**C)**. Expression values are compared between METABRIC discovery and validation batches within PAM50 molecular subtype classifications, allowing visualisation of potential batch correction signal augmentation. Standard non-parametric and parametric correction appear worse at minimising between-batch differences within each PAM50 molecular subtype. The addition of PAM50 molecular subtype as a covariate can improve correction while a stratified PAM50 approach removes batch effects most effectively. Pairwise comparisons between batches were performed with a two-sided Mann-Whitney-Wilcoxon test followed by Holm-Bonferroni correction. Non-significant (p > .05) comparisons are unlabelled while * denotes p <= .05, ** denotes p <= .01, *** denotes p <= .001 and **** denotes p <= .0001.

The third and final biomarker evaluated is expression of erbb2 receptor tyrosine kinase 2 (ERBB2). Significant expression difference between batches are introduced to HER2 PAM50 samples by parametric (p=.011) and non-parametric (p=.014) correction. Luminal A samples have between-batch significance removed by all correction techniques except PAM50 covariate correction (p=.046).

Leveraging the METABRIC discovery and validation datasets, we have outlined the importance of considering molecular subtype when batch-correcting before dataset integration. An additional interesting scenario arises when an individual dataset is produced at various time-points or by multiple laboratories, introducing potential batch effect manifestation. To investigate this premise, we consider a within-dataset evaluation of GSE6532^28^, a published dataset used in multiple studies^29^–31 that has been identified as displaying technical batch effects^32^. Samples within GSE6532 were ComBat corrected using the four approaches previously described.

Following batch correction, augmentation of GSE6532 ESR1 expression (figure 2) displays a similar outcome to METABRIC correction, with fewer significant differences following PAM50 covariate and stratified approaches. Basal samples, located only in batches one and two within this dataset, have significant population expression differences introduced following parametric (p=.006) and non-parametric (p=.022) correction. Luminal B samples display significant population differences following parametric correction (batches 2&3, p=.024 and batches 1&3, p=.024), non-parametric (between batches 2&3, p=.011 and 1&3, p=.011) and PAM50 covariate ComBat (batches 2&3, p=.05), with only PAM50 stratified correction displaying no significance between corrected batches. Normal-like samples continue to display significant differences following parametric (batches 1&3, p<.001 and batches 1&3, p=.014) and non-parametric (batches 1&3, p<.001 and batches 2&3, p=.023) ComBat.

**Figure 2.**
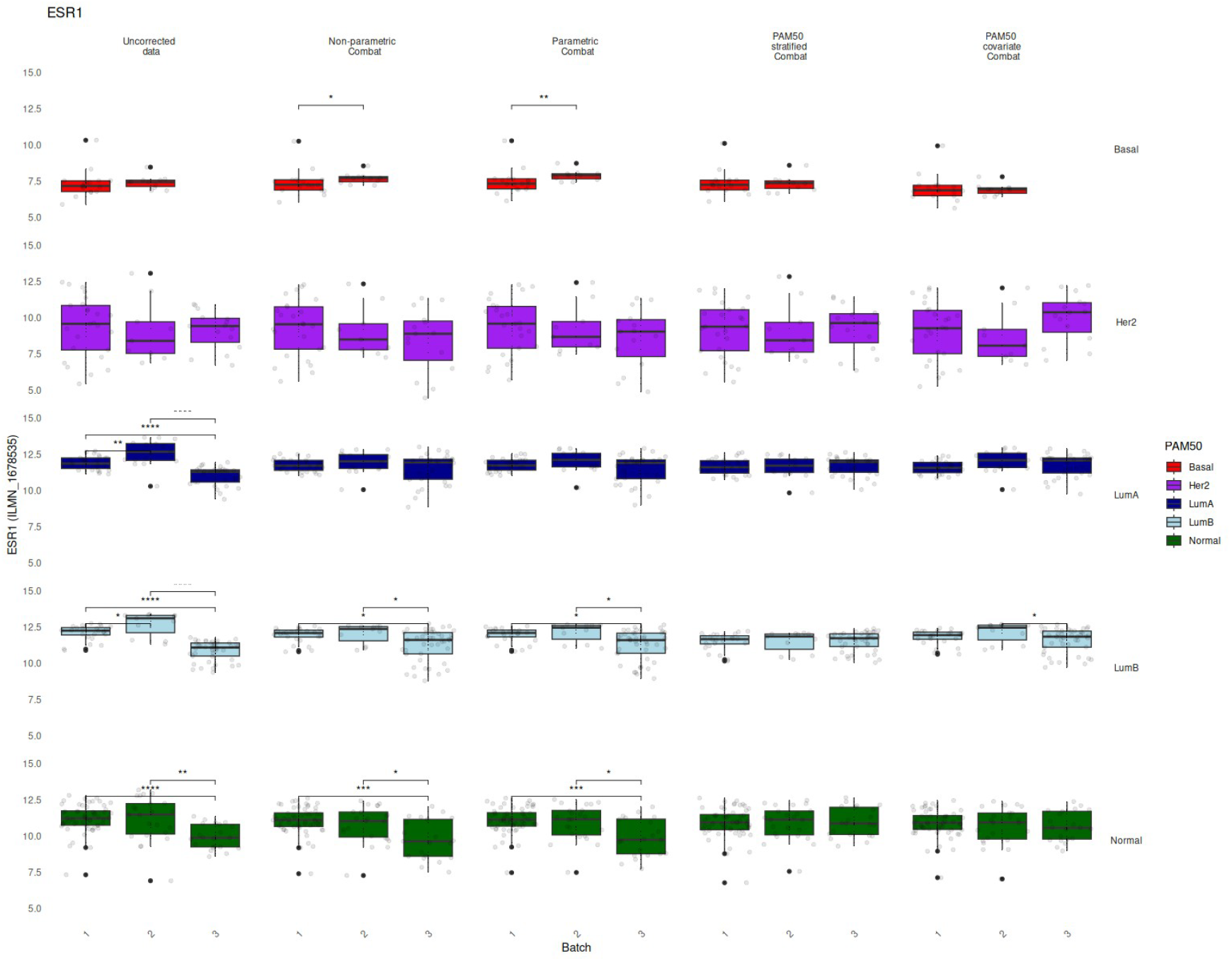
Effect of various ComBat batch correction implementations on GSE6532 ESR1 biomarker expression. Comparison of sample ESR1 expression values before and after 5 batch corrections. Samples were identified as belonging to three batches based on original sample processing date and stratified according to PAM50 molecular subtype class. Expression values are compared between batches within each subtype class following batch correction to visualise correction effects. ComBat non-parametric and parametric correction introduces significant signal difference between batches in Basal samples and performs poorly in Luminal B and Normal samples. Recognition of molecular subtype improves performance, with PAM50 stratified correction removing batch effects most effectively. Pairwise comparisons between patient groups were performed with a two-sided Mann-Whitney-Wilcoxon test followed by Holm-Bonferroni correction.Non-significant (p > .05) comparisons are unlabelled while * denotes p <= .05, ** denotes p <= .01, *** denotes p <= .001 and **** denotes p <= .0001.

The data presented herein illustrate often underappreciated subtleties in batch effect correction. Despite being a vital prerequisite for powerful data integration analysis, undesirable augmentation of molecular signal has the potential to seriously impact integrated analysis. Departures in molecular expression measurements between batches can be reasoned as composites of technical batch effects and biological, demonstrated by sample molecular subtype composition, structured differences. While the goal of batch correction is to address the former while retaining the latter, and acknowledging that no method will ever perfectly dissect these components, within this brief communication we highlight that additional consideration must be given to potential sub-optimal expression differences persisting following correction. The popular ComBat algorithm typically assumes identical distribution of gene expression across batches, but this assumption can be violated by divergent sample transcriptome profiles characteristic of molecularly heterogeneous cancers such as breast. To investigate this effect, we compared both standard parametric and non-parametric ComBat with PAM50-leveraged correction approaches. Our results show that encapsulation of molecular heterogeneity can help optimise the desired removal of technical effects while preserving true biological signal. It is perhaps reasonable and logical that incorporating molecular subtype classes within batch correction pipelines provide an effective shielding of molecular heterogeneity but, to our knowledge, this has not been elucidated until now. Within this article we purposefully refrain from definitively suggesting a particular batch correction approach. Rather, we invite consideration of an often overlooked, but potentially serious, feature of molecularly heterogeneous gene expression analysis that has the potential to seriously impact analysis and resulting scientific inference^33^.

## Data availability

METABRIC datasets are available via committee approval. GSE6532 is freely available at https://www.ncbi.nlm.nih.gov/geo.

## Additional Information

Our colleague Dr. Andrew H. Sims sadly passed away during the course of this project. He was instrumental in both the conceptualisation and supervision of this study.

